# *‘Parenting with Hope’* program among bereaved families in Colombia: A pre-post and quasi-experimental evaluation

**DOI:** 10.64898/2026.01.26.26344879

**Authors:** Andrés Villaveces, Sydney Tucker, Stiven Arroyo, Pedro José Blanco, Mayerlis Colón, Hilda E. Prías, Simona Pecsérke, Nicole Baldonado, Seth Flaxman, Susan D. Hillis, Oliver Ratmann

## Abstract

**Background:** In Colombia, internal violence, displacement, COVID-19, suicide, and climate crises threaten the survival of younger adults, many of whom are parents. Such premature mortality increases orphanhood risks. Evidence-based psychosocial support for surviving caregivers has potential to mitigate adverse impacts of orphanhood for bereaved children in crisis settings. Here, we adapted the *‘Hope Group’* program from war-affected Ukraine to post-COVID-19 Colombia *‘Parenting with Hope’,* and evaluated the effectiveness of psychosocial and parenting support delivered via home visits on improvements in caregiver mental health, violence against children, parenting practices, and child behavioral issues.

**Methods:** Participants (n=220) included surviving caregivers co-residing with children experiencing death of a parent or caregiver in the previous 12-36 months. Next-of-kin caregivers were identified through vital statistics data, death certificate annexes, radio/social media, schools, COVID-19 laboratories, and referrals. We used pre-post and quasi-experimental approaches to evaluate the effectiveness of *‘Parenting with Hope’*. For both analysis types, we constructed Bayesian models to estimate mean change and percent change following completion of the 8-session program.

**Results:** Both pre-post and quasi-experimental findings showed significant improvements across all mental health, violence prevention, parenting, and child outcomes. Pre-post results showed caregiver depression/anxiety ratings decreased by 91.2% (95% posterior credible interval (CrI) - 93.7, –87.6), and hopefulness increased by 43.8% (95% CrI 34.5, 54.8) and self-care, by 139.5% (95% CrI 107.5, 178.1). Each component measure of parenting practises (nonviolent discipline, positive parenting, parental monitoring, and parental involvement) improved significantly. By endline, violence against children had decreased by 63.9% (95% CrI -71.1, -54.4), and child externalizing and internalizing behaviors, by 74.4% (95% CrI, -78.0%, -70.3%). Pre-post and quasi-experimental findings showed equivalence.

**Conclusion:** This study generalizes evidence for effectiveness of *‘Parenting with Hope’* in crisis settings to surviving Colombian caregivers, on improved mental health, parenting practices, and reduced violence against children and child behavioral issues.

**Article Summary:** *‘Parenting with Hope’* for families in crisis settings significantly improved mental health and parenting strategies in post-COVID-19 Colombia, generalizing effectiveness previously measured in war-affected Ukraine.

**What’s Known on This Subject:** Caregiver death harms children long-term. Colombia experiences both community violence affecting caregivers and substantial COVID-19-related parental death burden. A Ukrainian program improved caregiver mental health, parenting, and reduced child violence, offering a model for Colombia’s prevention efforts.

**What This Study Adds:** We report pre-post evidence of the *Parenting with Hope’s* effectiveness in Colombian families experiencing bereavement. Adapted from Ukraine *‘Hope Groups’*, the intervention shows consistent benefits and is a promising, transferable and scalable strategy to prevent violence against children in communities globally.

## Background

Over one half of the 48 million people who live in Colombia are between ages 15-44 years old, according to 2025 data.^1^ In Colombia during the past two decades, escalating threats to the wellbeing and survival of younger adult populations, many of whom are parents and caregivers, include community violence, interpersonal violence, conflict, displacement, suicide, COVID-19 and other diseases, and climate hazards. A number of these threats are interrelated and linked to social determinants of health, including multi-dimensional poverty.

As an example of the interconnectedness of such threats, community violence has led to large proportions of the population being internally displaced — about 7.3 million persons by 2024.^2^ Such internal displacement can further threaten parents and children, through family disruption, separation, and premature death. For males, the second top cause of death is violence,^3^ and deaths due to violence and suicide occur more commonly among younger adults.^3^ In recent years, Colombia also experienced high mortality of caregivers during the COVID-19 pandemic, leaving an estimated 55,000 children without parents or caregiver support.^4^ Intersecting with these multiple causes of mortality is widespread poverty, as approximately 36% of the total population and 48% of children under 15 live below the poverty line.^5^ Colombia further has one of the highest global levels of social inequalities, ranking fifth worldwide, with a GINI coefficient of 53.9 for 2023.^6^ Colombia also ranks 10^th^ globally in terms of economic risk posed by climate-related hazards.^7^ The combined impacts of multi-dimensional poverty and early deaths of parents or caregivers have long-term health consequences for surviving family members and children, including negative mental health outcomes^8,9^ increased risks of further violence perpetration,^10^ and increased risks of sexual victimization and exploitation.^11,12^

Evidence suggests that children can be protected from adverse consequences of caregiver death through interventions for surviving caregivers that improve parenting skills,^13^ strengthen caregiver mental health,^14,15^ build relationships between parents and children, and strengthen violence-prevention skills.^16^ Such interventions build resilience among children and their families and reduce risks of caregiver depression and anxiety, violence against children, and protect children from sexual abuse and exploitation.^17,18^ Thus, we aimed to: 1) evaluate options for identifying caregivers for children experiencing bereavement, including through vital statistics data, death certificate annexes, radio/social media, and schools;^19,20^ 2) evaluate the effectiveness of the adapted parenting intervention *‘Parenting with Hope’* from the Ukrainian *‘Hope Groups’* using distinct analytic approaches; 3) assess bereaved families to refer them to needed services; and 4) compare intervention effectiveness between the Colombian post-COVID-19 and Ukrainian war-time settings.

We selected a region affected by the previously described vulnerabilities within the Department of Sucre, Northern Colombia. To identify eligible caregivers for children who had experienced orphanhood, we extended methods previously described, including the use of a death certificate field listing child dependents in the home.^19,20^ We adapted the *‘Hope Groups’* parenting program^17^ (initially implemented in Ukraine with families displaced by war), to the Colombian orphanhood context and called the adapted version *‘Parenting with Hope’*. Salient program characteristics include community participation, activities that improve parent-child communication, and community-based delivery of the intervention, given strong evidence of effectiveness for such programs.^21^ This approach aligns with the National Action Plan to prevent violence against children^22^ which identified parenting interventions as a key priority in response to findings from the 2018 VAC survey.^11^ Research in Colombia shows that reducing stigma about mental health^14^, improving communication within families^15^ and linking families to services^23^ are the most needed strategies to strengthen family wellbeing among vulnerable among vulnerable children and youth. The intervention was implemented with participation of psychologists and social workers working with our local partners from the University of Sucre who have extensive experience of work within the communities of the Department of Sucre.

Implementation of *‘Parenting with Hope’* was designed as a semi-structured psychosocial and parenting support group delivered face-to-face at the home. It consisted of 8 parenting sessions (two less than Ukraine *‘Hope Groups’* because it was done outside a war context). Absence of caregivers due to any cause of death was a key eligibility criterion for identifying families.^24^ The intervention included mental health content and used psychosocial support principles to strengthen or reorient parental skills to improve resilience through building coping approaches, stress reduction, self-care, and understanding of child behavior during times of crisis.^24^ Other contents including caregiving and violence prevention drew from global programs such as Parenting for Lifelong Health (PLH) at the University of Oxford, an intervention endorsed by WHO and with the addition of evidence-based gender-based violence components for the family.^25,26^ The contents of the adapted intervention were also reviewed by the creators of the *‘Hope Groups’* for fidelity. The delivery of sessions was planned according to family preferences and availability (typically once a week) for about 90 minutes. Each participant received a copy of the *‘Parenting with Hope’* guide (WWO).

For this study we hypothesized that participation in these sessions with bereaved families who had recently lost a caregiver, including those who were further affected by social and political contexts in Colombia, would improve one or more outcomes of positive parenting, mental health, prevention of violence against children in the home and improved family and child wellbeing.^27^ Grief experienced by families due to caregiver death, who at times were further threatened by exposures to violence or displacement, is a crucial trauma to address, to prevent negative consequences of orphanhood and other adverse childhood experiences. *‘Parenting with Hope’* strengthens psychosocial support for adults to build participants’ mental health skills in healthy coping and grieving, self-care for themselves and their children, stress reduction, and empathetic understanding of a child’s behavior under these conditions. The *‘Parenting with Hope’* strategies focus on reinforcing positive behavior, supporting child development, child supervision and protection, and use of communication and non-violent discipline in compliance with Colombian legislation addressing the prohibition of all forms of violent and degrading behaviors against children, Law 2089 of May 2021.^28^ Given initial positive findings from the Ukraine *‘Hope Groups’* in a war context,^17^ this study evaluates the effectiveness of our adapted *‘Parenting with Hope’* program, using similar outcomes for caregivers of children affected by loss of parents and caregivers in an intersecting crises context of poverty, community violence and ensuing impact of COVID-19 parental deaths. We thus aim to demonstrate external validity of the intervention and provide evidence for policies that scale and sustain *‘Parenting with Hope’/‘Hope Groups’* in Colombia and beyond.

## Methods

### Study design

We used pre-post and quasi-experimental statistical designs to obtain evidence on the feasibility and effectiveness of *‘Hope Groups’* adaptation in Colombia. Given the crisis setting, we initially selected a pre-post design for its feasibility over a randomized design. Because our intervention rolled out to participant cohorts over time, we were additionally able to conduct a quasi-experimental analysis to strengthen the rigor of our results by accounting for time trends. This quasi-experimental design relies on principles from rolled-entry matching,^29^ where participants completing a treatment are compared with participants starting a treatment, and incidence-density case-control sampling,^30^ where participants with a specific condition are matched with participants without the condition at the same calendar timepoint. In our study, we compared cohorts of participants completing the intervention with new participant cohorts who were enrolling in the intervention at the same calendar timepoint. Specifically, we compared outcomes of participants completing their endline surveys with the outcomes of participants who were completing their baseline surveys within two weeks of each other. The primary independent variable was the 8-session *‘Hope Group’* program, adapted for Colombia under the name *‘Parenting with Hope’*, and the primary outcomes included caregiver mental health, parenting practices, and violence against children. Modeling accounted for key potential confounders such as time since caregiver death.

### Setting

This study was conducted in Northern Colombia (Department of Sucre), among families affected by bereavement due to any cause. Participants were recruited from next-of-kin caregivers of bereaved children, defined as children who had experienced the death of a primary caregiver. The intervention was delivered in urban and peri-urban communities of five municipalities (Sincelejo, Corozal, San Onofre, Sampués, San Marcos), hrough either individual home visits or small micro-group sessions (2-4 participants). All sessions followed a standardized protocol structure and adhered to strict confidentiality agreements. This flexible delivery model facilitated the inclusion of diverse family structures and socioeconomic backgrounds while maintaining rigorous participant protection standards and ensuring data collection integrity throughout the study period.

### Intervention adaptation and contextualization, training, recruitment, and implementation

We used the ADAPT guidance to systematically adapt the *‘Hope Groups’* model, previously validated among Ukrainian caregivers amidst active war during the Russian invasion,^17^ for surviving Colombian caregivers of orphaned children. The localization process, led by subject matter experts affiliated with the University of Sucre, involved a structured, multi-week protocol which encompassed translation, cultural contextualization, and content re-sequencing to address bereavement and crisis-related stressors, rather than active conflict scenarios. The delivery modality for the Colombian *‘Parenting with Hope’* program converted the original group-based *‘Hope Groups’* format to a flexible home-visit model, accommodating individual caregivers or micro-groups of 2-4 family members. Core safeguarding protocols were maintained and enhanced, including strict confidentiality measures, clear delineation of facilitators’ non-therapeutic roles, and established mental health referral pathways for participants requiring clinical intervention. The underlying *‘Hope Groups’* original theoretical framework and evaluation methodology, including standardized pre- and post-intervention assessments, remained unchanged to preserve intervention fidelity and enable cross-cultural outcome comparisons across diverse contexts.

Colombian facilitators received targeted training on the adapted *‘Parenting with Hope’* Guide, which covered session structure, integration of at-home practice activities, and implementation of safeguarding protocols, including distress recognition, mandatory reporting, and referral pathways. Facilitators were also trained in standardized pre- and post-intervention data collection procedures, including simulated practice sessions to reinforce delivery skills. Regular supervisory checkpoints were established to clarify escalation criteria and ensure adherence to consistent, trauma-informed communication within home-based intervention settings.

Next-of-kin caregiver participants were recruited through a systematic, multi-source sampling strategy designed to maximize population reach and minimize selection bias. Recruitment sources for surviving parents and caregivers included: municipal mortality registries from Sincelejo (Capital of Sucre), and the four additional participating municipalities; targeted social media and radio campaigns; referrals from public educational institutions; partnerships with local funeral service providers; COVID-19 testing laboratory databases; and word-of-mouth referrals from other bereaved families. All potential participant records underwent systematic de-duplication procedures and eligibility verification to confirm bereavement status and primary caregiver responsibilities. Initial contact was established using standardized, trauma informed communication protocols developed specifically for bereaved populations. Home-based recruitment visits were conducted to ensure equitable access across socioeconomic strata and geographical locations. Informed consent and assent procedures followed standardized protocols regardless of recruitment source, maintaining methodological consistency and ethical compliance throughout the enrollment process.

The intervention was delivered primarily through home-based visits, with participants permitted to include family members, neighbors, or additional kinship caregivers to form micro-groups when appropriate. All sessions adhered to a standardized protocol structure comprising opening agreements, practice of previously introduced coping strategies, and guided reflection activities. The intervention explicitly avoided trauma narrative exposure; instead, facilitators employed trauma-informed approaches that prioritized participant safety and emotional regulation. The implementation model was designed for scalability and replicability, incorporating minimal resource requirements, clearly defined facilitator role boundaries, flexible delivery modalities, and a streamlined risk management framework encompassing comprehensive safeguarding measures and mandatory reporting procedures. Implementation fidelity and data integrity were maintained through systematic quality assurance mechanisms, including weekly supervision sessions, structured case review process, validated fidelity assessment checklists, and standardized pre- and post-intervention outcome measurements. This multi-layered supervision structure ensured consistent intervention delivery while maintaining participant safety and research validity across all implementation sites.

### Data Collection Procedures

Imperial College London Ethics Committee and University of Sucre Ethics Committee approved this study. We used mortality databases and death certificate annexes, and other referral sources to identify potentially eligible next-of-kin caregivers for children after prior death of a co-residing parent or grandparent caregiver. We collected baseline and endline data via participant surveys administered by program staff. Informed consent was obtained prior to completion of baseline surveys. Informed consent and baseline surveys were administered via Kobo Toolbox before the first *‘Parenting with Hope’* session. Endline survey data were collected 2 weeks after completion of the 8-session *‘Parenting with Hope’*, also via Kobo. To reduce social desirability bias, endline surveys were administered during home visits from another trained program staff member, and never by a participant’s *‘Parenting with Hope’* facilitator. No incentives were offered for survey completion. All data were pseudonymous – utilizing only unique identifiers created from initials and birth month, instead of full names. Our protocol required immediate referrals to child protection and health services in the event any participant reported severe abuse, rape, or other significant harm during survey completion sessions or *‘Parenting with Hope’* sessions. However, no such referrals were needed.

### Outcomes

We adapted standardized and validated primary and secondary outcomes from the original *‘Hope Groups’* study among Ukrainians impacted by war, as previously described.^17^ For our caregiver mental health primary outcome, we selected <u>the 6-item Kessler scale</u> used in Violence Against Children surveys globally^31^ to measure depression and anxiety, utilizing a likert-frequency scale ranging from ‘None of the time’ to ‘All of the time’. For all remaining outcomes, we used frequency scales to assess mean change of occurrence in ‘days in the last week’ to strengthen recall for surviving caregivers having experienced bereavement, as done in Ukraine.^17^ We further measured, as primary outcomes: violence against children using behavioral indicators from the International Society for Prevention of Child Abuse and Neglect Screening Tool (I-CAST-Trial tool for parents – Spanish version),^32^ and parenting practices via indicators from the Alabama Parenting Questionnaire (including nonviolent discipline, positive parenting, parental monitoring, and parental involvement).^33^ As secondary outcomes, we measured child wellbeing using items from the Child and Adolescent Behavior Inventory (internalizing and externalizing behaviors), and caregiver well-being outcomes including hopefulness about the future and self-care practices.^34^

### Sample Size Calculation

Sample size calculations were conducted using the pwr package in R (v 1.3-0), based on estimated effect sizes and variance parameters from the *‘Hope Groups’* evaluation in Ukraine.^17^ The study was powered to detect a statistically significant absolute reduction in violence against children. Estimating an average cluster size of six and 20% loss-to-follow-up, our target sample size was 240.

### Data Cleaning and missingness

All data were cleaned and analyzed in R (version 2024.04.2). Missingness was low; 91.6% of participants who completed a baseline survey completed an endline survey, and demographic and outcome variables were rarely missing (<1%).

### Analytic Approach

#### Pre-post analysis

We constructed Bayesian partial credit models^35^ to estimate mean change and percent change associated with completion of 8-session *‘Parenting with Hope’* for 220 participants from ordered categorical data. The main model included a single latent factor for each participant to capture their latent trait, a fixed intervention effect that modulated baseline latent trait, response item loadings to capture how participant traits map to measured behaviors at baseline and endline, and partial credit thresholds that encode the challenge in attaining the next category in likert-scale or day-of-week outcomes. Additional adjusted models included a fixed effect on a key confounder of time since caregiver death (in months), and a propensity score including numerous other potential confounders (age, sex, food insecurity, number of adults and children in household, child disability, education, income, marital status, and receipt of government resources). The models were fitted with the Stan probabilistic computing language,^36^ and statistical uncertainty was quantified using 95% credible intervals (CrI) in posterior densities. We used posterior predictive checks to validate the model (Supplementary Figs. S1, S2), and found very good concordance across all item responses, improving on linear models.^17^ We tested the partial credit models against alternative item-response models including the ordered logistic model^37^ using importance sampling approximations to leave-one-out cross-validation.^38^ The ordered logistic model had similar posterior predictive check and cross-validation characteristics as the partial credit model, but numerical inference was by far most stable for the partial credit model. We also compared the pre-post results in Colombia with the pre-post results in Ukraine.^17^

### Quasi-experimental analysis

We used the MatchIt package in R (v. 4.7.0) to match participants completing their endline surveys with participants completing baseline surveys at the nearest calendar timepoint (within a maximum allowed range of 2 weeks). Any baseline surveys which did not align with endline surveys were excluded, as were endline surveys which did not align with baseline surveys. Analysis included 121 baseline and endline surveys. We used the same Bayesian partial credit model as in the Pre-post analysis, adding a fixed effect on the matching strata. We additionally investigated time since caregiver death with calendar time to ensure there were no positivity violations (Supplementary Figure S3). Statistical uncertainty was also quantified via 95% credible intervals. As a sensitivity analysis, we compared results with a standard frequentist approach using a two-level generalized mixed effects model, and results were consistent.

## Results

We examined characteristics of the study population including caregiver age, education, income, municipality, household size, food insecurity, and select bereavement indicators. We found that the majority of caregivers were ages 20-60 (87%), female (92%), lived in Sincelejo (78%), resided in households with 2 or more caregivers (81%), had a median of 2 bereaved children living in the home, had a household monthly income of 0-500,000 pesos (equaling $0-$135 US dollars), and reported anxiety about hunger (97%) (Table 1). Similar distributions were observed for marital status (married/partnered (35%) and single/separated (38%); approximately 1/4 (26%) reported having up to primary school education, with the remainder reporting secondary school (49%) or higher (25%).

**Table 1:** Bereaved Next-of-Kin Caregivers’ Baseline Characteristics in Colombia (2024), Categorized by Time Since Bereavement.

|  | Less than 1-year post-bereavement | More than 1-year post-bereavement | Overall |
| --- | --- | --- | --- |
|  | (N=141) | (N=99) | (N=240) |
| **Caregiver Age** |  |  |  |
| Less than 20 | 4 (2.8%) | 1 (1.0%) | 5 (2.1%) |
| 20-30 | 18 (12.8%) | 21 (21.2%) | 39 (16.3%) |
| 30-40 | 42 (29.8%) | 25 (25.3%) | 67 (27.9%) |
| 40-50 | 28 (19.9%) | 21 (21.2%) | 49 (20.4%) |
| 50-60 | 29 (20.6%) | 19 (19.2%) | 48 (20.0%) |
| 60+ | 20 (14.2%) | 12 (12.1%) | 32 (13.3%) |
| **Sex** |  |  |  |
| Female | 135 (95.7%) | 91 (91.9%) | 226 (94.2%) |
| Male | 6 (4.3%) | 8 (8.1%) | 14 (5.8%) |
| **Marital Status** |  |  |  |
| Divorced | 1 (0.7%) | 1 (1.0%) | 2 (0.8%) |
| Living with partner | 44 (31.2%) | 29 (29.3%) | 73 (30.4%) |
| Married | 14 (9.9%) | 7 (7.1%) | 21 (8.8%) |
| Separated | 10 (7.1%) | 4 (4.0%) | 14 (5.8%) |
| Single | 40 (28.4%) | 34 (34.3%) | 74 (30.8%) |
| **Education** |  |  |  |
| Less than Primary School | 6 (4.3%) | 5 (5.1%) | 11 (4.6%) |
| Primary School | 36 (25.5%) | 21 (21.2%) | 57 (23.8%) |
| Secondary School | 60 (42.6%) | 48 (48.5%) | 108 (45.0%) |
| Technical School | 30 (21.3%) | 19 (19.2%) | 49 (20.4%) |
| University | 7 (5.0%) | 6 (6.1%) | 13 (5.4%) |
| Missing | 2 (1.4%) | 0 (0%) | 2 (0.8%) |
| **Number of Adults in Household** |  |  |  |
| Single Caregiver Household | 22 (15.6%) | 19 (19.2%) | 41 (17.1%) |
| 2 Adults in Household | 48 (34.0%) | 35 (35.4%) | 83 (34.6%) |
| 3+ Adults in Household | 71 (50.4%) | 45 (45.5%) | 116 (48.3%) |
| **Household Income** |  |  |  |
| 0-500 | 93 (66.0%) | 67 (67.7%) | 160 (66.7%) |
| 500-1M | 30 (21.3%) | 21 (21.2%) | 51 (21.3%) |
| 1M-1.5M | 10 (7.1%) | 7 (7.1%) | 17 (7.1%) |
| 1.5M-2M | 4 (2.8%) | 3 (3.0%) | 7 (2.9%) |
| >2M | 4 (2.8%) | 1 (1.0%) | 5 (2.1%) |
| **Number of Children in Household** |  |  |  |
| Mean (SD) | 2.11 (1.29) | 2.26 (1.18) | 2.17 (1.25) |
| Median [Min, Max] | 2.00 [1.00, 10.0] | 2.00 [1.00, 7.00] | 2.00 [1.00, 10.0] |
| **Number of Bereaved Children in Household** |  |  |  |
| Mean (SD) | 1.99 (1.08) | 2.13 (1.14) | 2.05 (1.10) |
| Median [Min, Max] | 2.00 [1.00, 6.00] | 2.00 [0, 6.00] | 2.00 [0, 6.00] |
| **Household Municipality** |  |  |  |
| Corozal | 6 (4.3%) | 4 (4.0%) | 10 (4.2%) |
| Sampués | 1 (0.7%) | 2 (2.0%) | 3 (1.3%) |
| San Marcos | 5 (3.5%) | 6 (6.1%) | 11 (4.6%) |
| San Onofre | 12 (8.5%) | 10 (10.1%) | 22 (9.2%) |
| Sincelejo | 117 (83.0%) | 77 (77.8%) | 194 (80.8%) |
| **Experiencing Food Insecurity** |  |  |  |
| Yes | 137 (97.2%) | 97 (98.0%) | 234 (97.5%) |
| No | 4 (2.8%) | 2 (2.0%) | 6 (2.5%) |

We used a number of recruitment strategies to identify bereaved adults living in households as surviving caregivers for bereaved children, including Sincelejo municipality mortality database, Sampués municipality mortality database, social media and radio announcements, school referrals, funeral home annex forms, COVID-19 laboratories, and referrals from other bereaved families, to identify deceased adults whose next-of-kin could identify surviving parents and caregivers for orphaned children. Upon examining the cascade from potential identification of adult deaths to the enrollment of surviving caregivers of bereaved children, we found that among 2862 deceased adults, 52.5% (1503) had available phone numbers, of which 45.9%% (690) could be reached, and 36% (249) of these next of kin reported orphaned children living in the household. Importantly, among next-of-kin identified, nearly all (97.2%, N=242) elected to enroll in the *‘Parenting with Hope’* program (Figure 1A). Upon further examination, the most effective recruitment strategies for identifying bereaved next-of-kin caregivers included social media/radio and Sincelejo mortality database, which together accounted for 66.1% (160) of enrolled households (Figure 1B). The number of children with potential to benefit from the *‘Parenting with Hope’* program steadily grew over the 57-week enrollment period (Fig 1C), as the number of children (518) across households was almost twice the number of caregivers enrolled (242).

**Figure 1.**
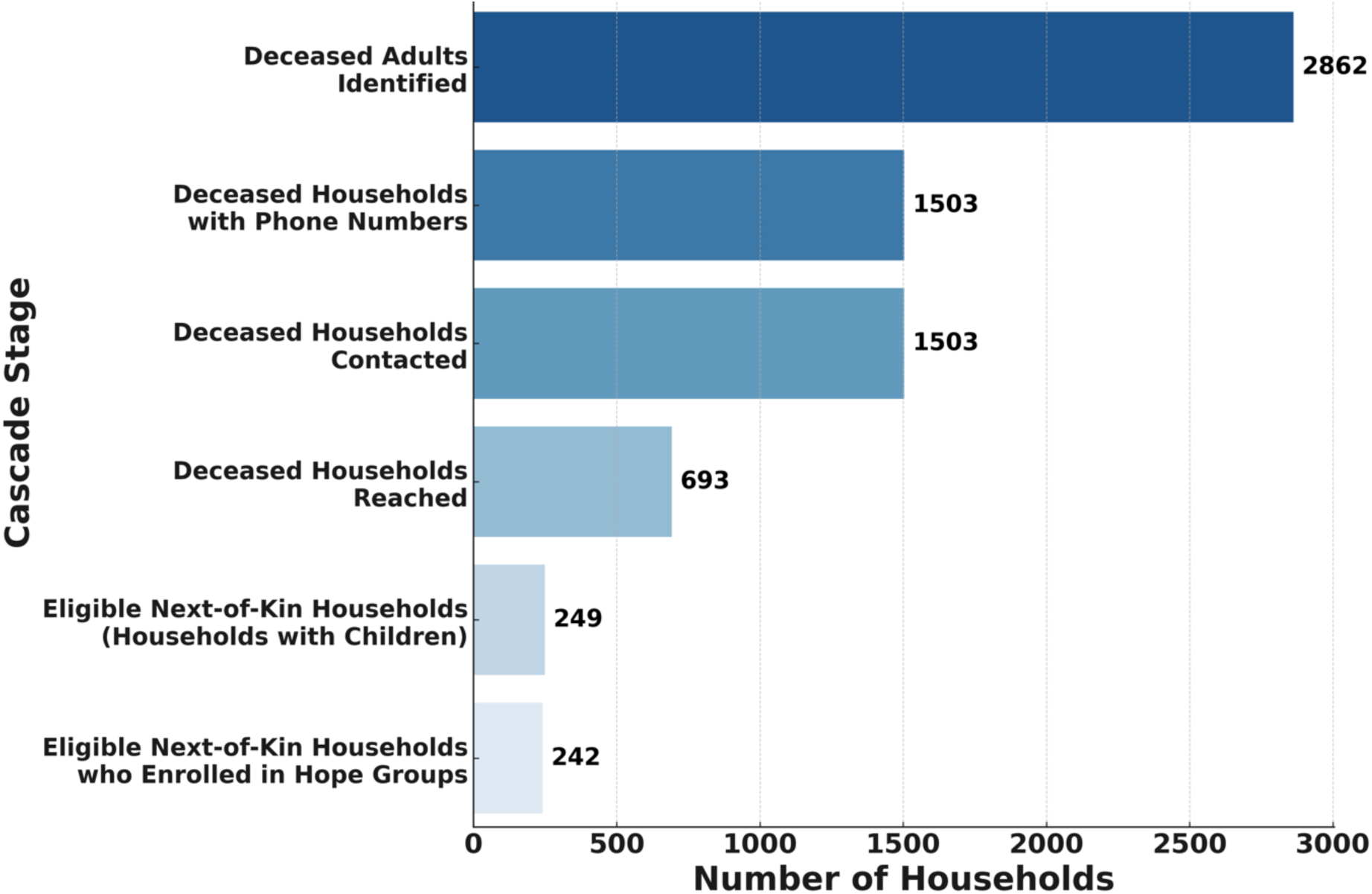
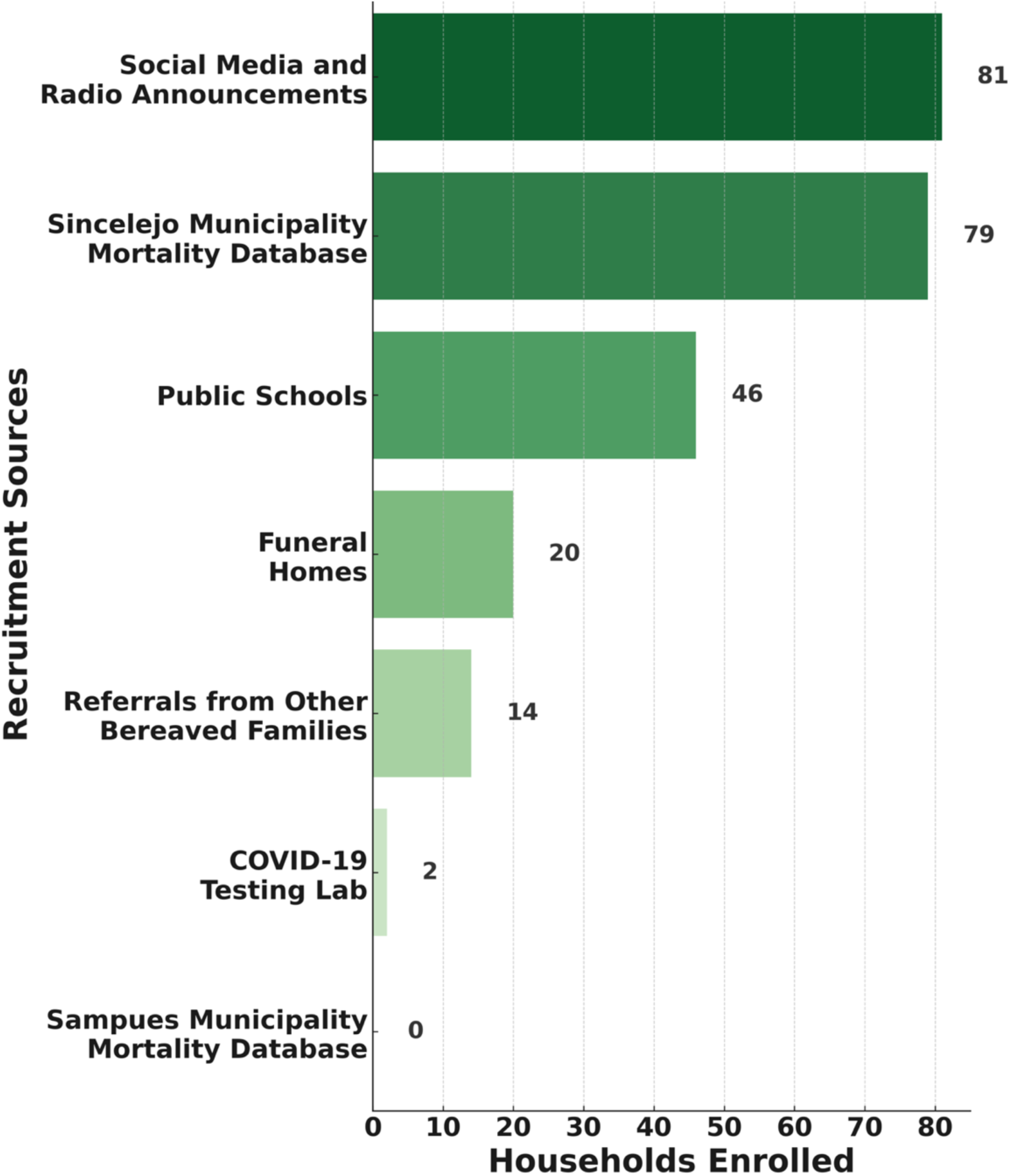
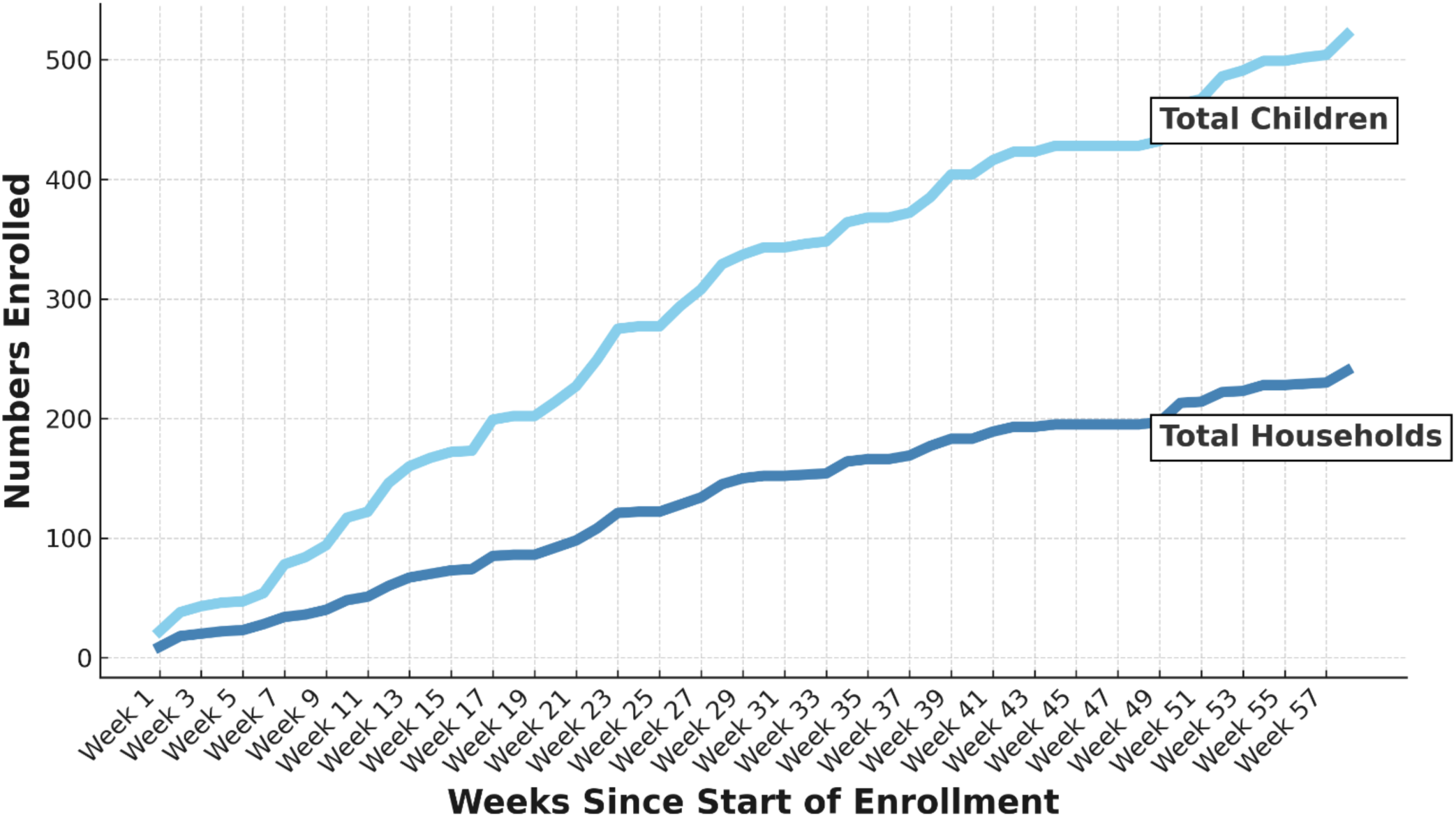
Identification of Deceased Caregivers and Recruitment of Next-of-Kin Caregivers into Family Strengthening Programming in Colombia (2024) A. Cascade to Care: The Process from Identification of Deceased Adults to Enrolling Bereaved Next-of-Kin Caregivers in Family Strengthening Programming B. Effective Recruitment Sources: Key Recruitment Sources to Enroll Bereaved Next-of-Kin Caregivers in Family Strengthening Programming in Colombia (2024) C. Recruitment Over Time: Recruitment of Bereaved Next-of-Kin Families in Colombia from February 2024 - December 2024 into ‘Parenting with Hope’, a Mental Health, Psychosocial, Parenting Strengthening Program

Our analysis of the 8-week *‘Parenting with Hope’* program found statistically significant improvements across all mental health, violence against children, parenting, and child primary and secondary outcomes when examining change from baseline-to-endline (Fig 2, Table 2). By endline, caregiver anxiety and depression, violence against children, and child behavioral issues had decreased, and significant improvements were observed in caregiver self-care and hope; as well as in parental involvement and monitoring, nonviolent discipline, and positive parenting (Fig 2). We found reductions in depression and anxiety ratings of 91.2% (95% CrI -93.7, –87.6), with reductions not driven by a single symptom, as improvements were observed across hopelessness, perceived effort, and worthlessness (Tables S1, S2). Program participation further was associated with increases in hopefulness, and self-care of 43.8% (95% CrI 34.5, 54.8) and 139.5% (95% CrI 107.5, 178.1), respectively (Table 2).

**Figure 2.**
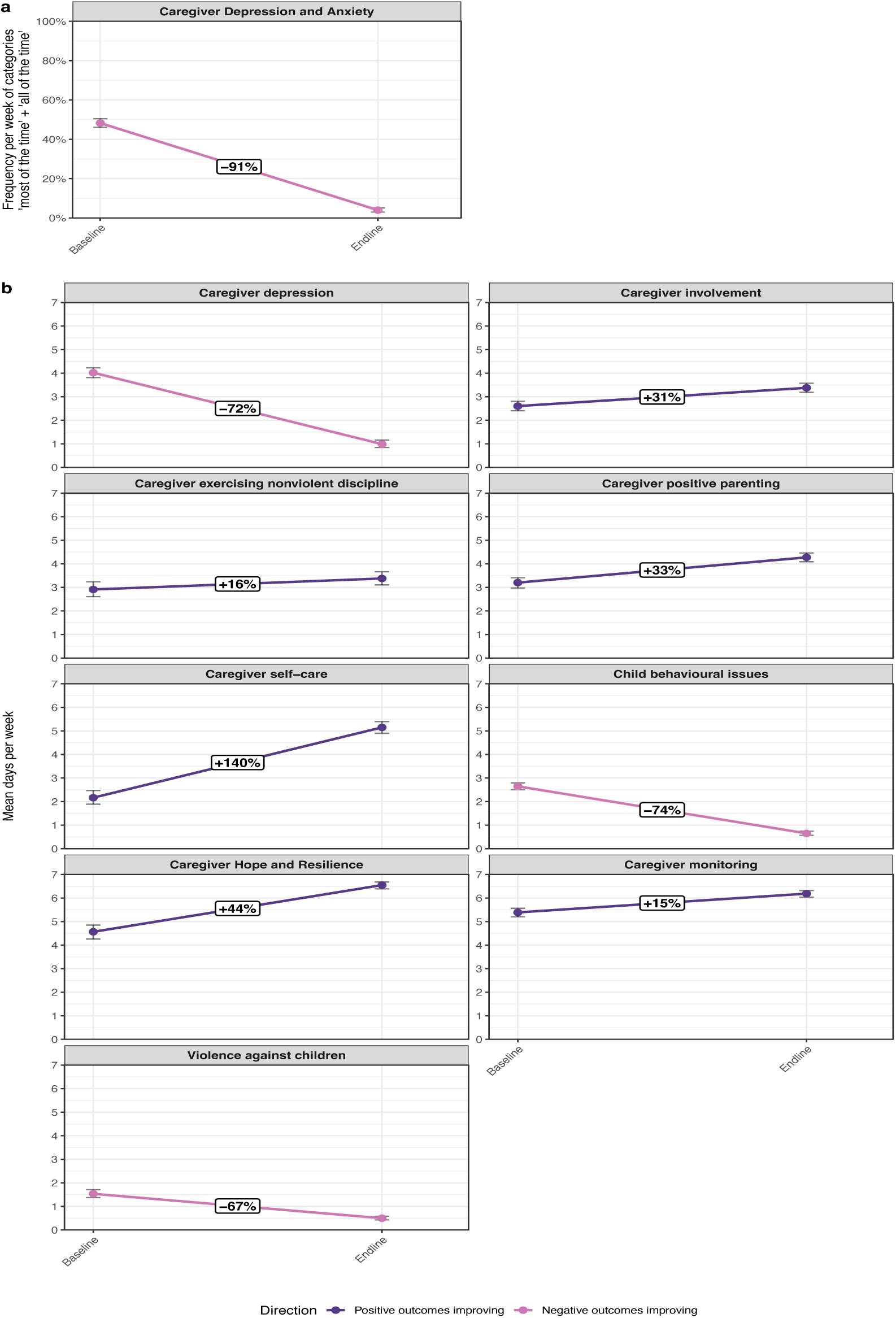
Change in Outcomes ‘Pre’ and ‘Post’ Participation in *‘Parenting with Hope’*, a Psychosocial, Mental Health, and Parenting Strengthening Program for Bereaved Next-of-Kin Caregivers in Colombia (2024-2025).

**Table 2:**
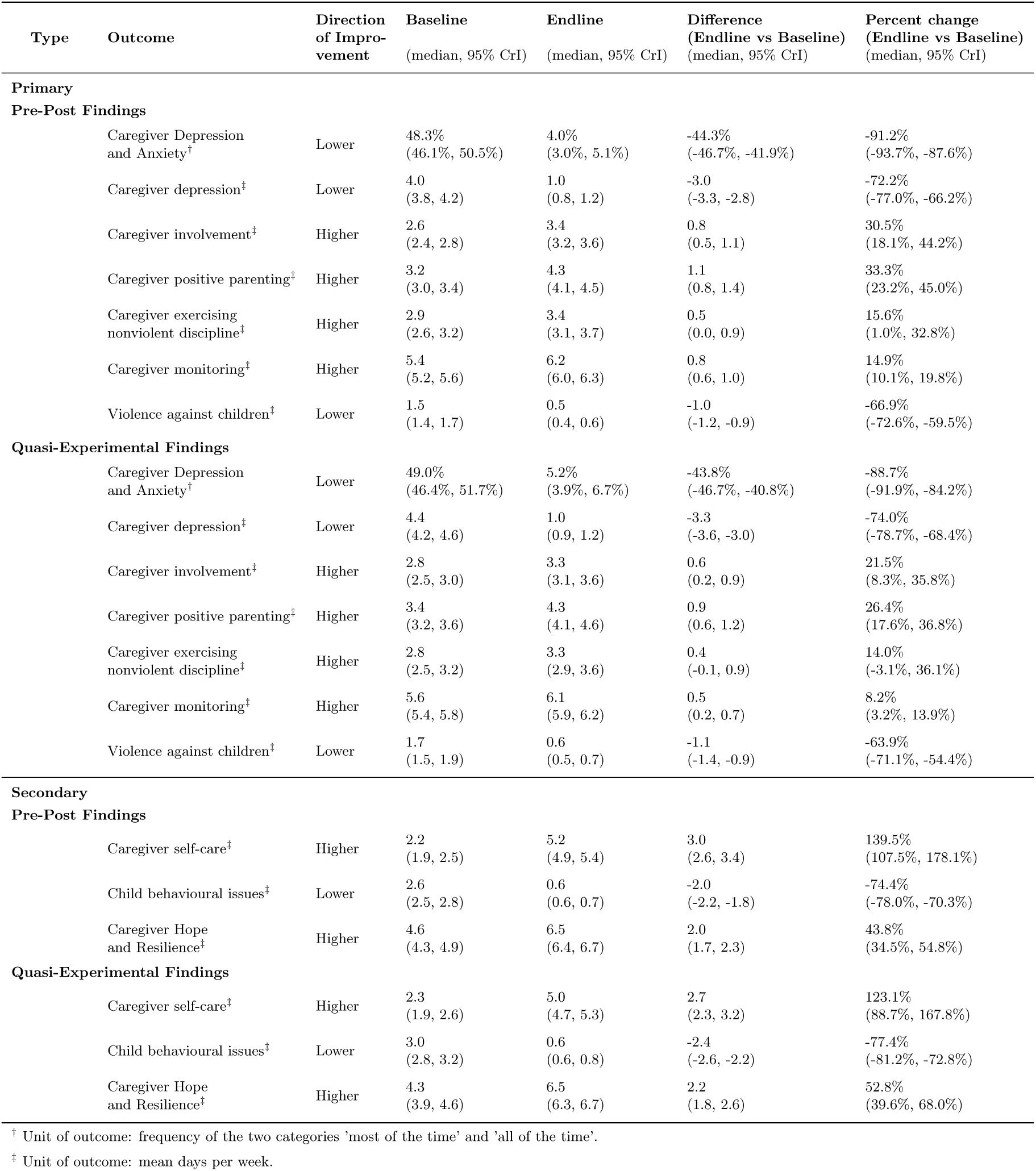
Changes in outcomes following Parenting with Hope participation among next-of-kin care-givers in Colombia in 2024-2025, utilising pre-post and quasi-experimental methodologies.

Results of our pre-post analysis showed that violence against children decreased by 63.9% by endline (95% CrI -71.1, -54.4), child behavioral issues decreased by 74.4% (95% CrI, -78.0%, - 70.3%) (Table 2), and all parenting practices improved, with increases in nonviolent discipline by 15.6% (95% CrI 1.0%, 32.8%), positive parenting by 33.3% (95% CrI 23.2%, 45.0%), monitoring child by 14.9% (95% CrI 10.1, 19.8); and parental involvement by 30.5% (95% CrI 18.1, 44.2). Effectiveness measures for each survey item are reported in Supplementary Table 1.

Importantly, based on our data for Colombia, results of our quasi-experimental analysis were consistent with those of our pre-post analysis (Table 2, Figure 3, Table S2). We further find that results comparing our findings for bereaved next-of-kin caregivers in Colombia with Ukrainian caregivers amidst war and displacement^17^ (Table S3) were strongly congruent. Thus, the equivalence of findings across the two analytic approaches for Colombia, along with the congruence between the Colombian and Ukrainian findings strengthens our conclusions regarding the substantial effectiveness of the *‘Parenting with Hope’* program for key mental health, violence prevention, practices, and child behavioral outcomes within two weeks after the end of the intervention.

**Figure 3:**
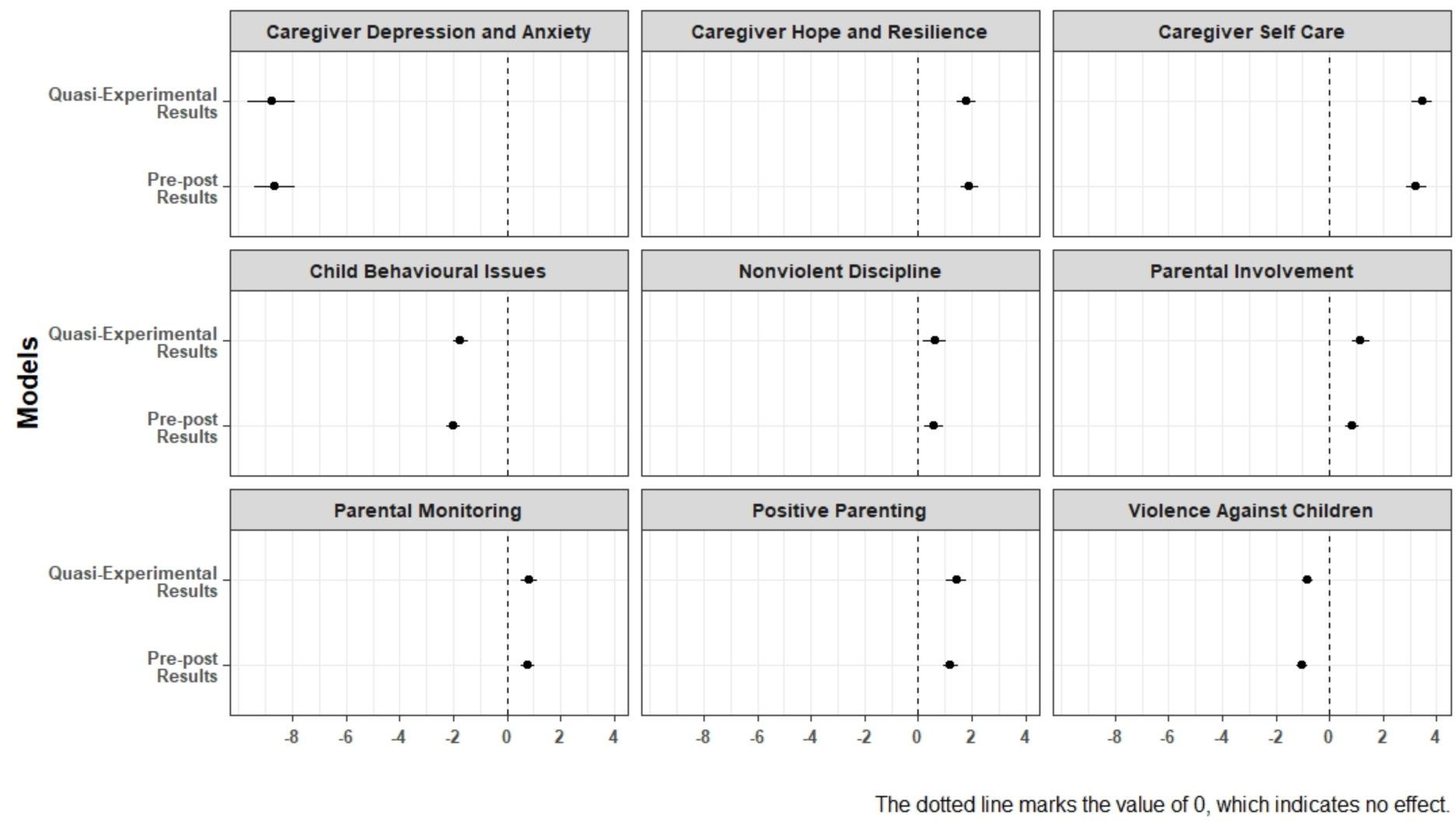
Comparing Results Across Pre-Post and Quasi-Experimental Stepped Entry Matching Methodologies, among Colombia Caregivers from 2024-2025.

## Discussion

Timely identification of children affected by orphanhood constitutes a key challenge for reaching their surviving caregivers with evidence-based psychosocial support programs to mitigate associated adverse impacts of parental death on children. We found a variety of strategies useful for facilitating timely identification of next-of-kin caregivers for such children, including municipal mortality databases, school referrals, and funeral home annexes. Almost all caregivers who were identified consented to enrollment in the *‘Parenting with Hope’* program. This 8-session intervention for bereaved caregivers of orphaned children led to significant reductions in caregiver anxiety and depression, improvements in caregiver resilience, self-care, and parenting practices, along with reductions in violence against children and child behavioral issues.

Strengthened caregiver mental health was shown by the 60% reduction in the frequency of participants’ anxiety and depression rating, over a 40% increase in hopefulness, and a160% improvement in self-care. Caregiver practice that helped protect children from violence, including nonviolent discipline, positive parenting, parental monitoring, and parental involvement, improved significantly. By study endline, violence against children decreased by nearly 70%, and child externalizing and internalizing behaviors, by 75%. We also found that nearly all participants reported food insecurity, and the *‘Parenting with Hope’* program linked participants to the Department of Social Protection, to strengthen economic support for participants.

Our findings indicate that the *‘Parenting with Hope’* program addresses a key public health gap in Colombia, as described in a systematic review showing that services in the country for mental health, families, and violence prevention are insufficient.^14^ The review further highlights the need to improve care in Colombia by prioritizing community participation, users’ perspectives, interinstitutional collaboration, and less reliance from biomedical models of care.^14^ The interinstitutional leadership of University of Sucre as the lead educational institution in-country for evaluating the program, has been key for sustainability of this program, as the Health Sciences Deans have integrated *‘Parenting with Hope’* into the curriculum, training upper level health sciences students to deliver the program as part of their practicums. Prompt implementation of interventions that are adapted for risks of bereaved children can restore hope and strengthen resilience, as confirmed by our findings. Conceptually, building parenting skills for surviving caregivers helps facilitate recovery for children by strengthening self-esteem, agency, and peer relationships.^4^ Previous studies confirm the complexity of social, economic, and psychological consequences of orphanhood, and the benefits of interventions that help children and their surviving caregivers find strength, experience growth, and develop new abilities.^39–41^ Extensive programming for orphans and vulnerable children such as those affected by pandemics have demonstrated that investments in evidence-based programs (e.g. cash transfers, parenting support and safe schools) promotes resilience, with benefits for children, families, communities, and nations.^42^ Thus, the care children receive after caregiver death shapes the consequences of that death.

In Colombia and around the world, policies and programs that protect children from the consequences of parental death, including violence, sexual exploitation, and mental health threats, are lagging^23^ at a time when risks for children are increasing due to escalating and coalescing crises, including conflict, climate, and epidemics. Because such crises threaten families and children through similar pathways – displacement and migration, parental death and illness, poverty and food insecurity, and disruption of basic services – it is feasible that a core crisis-adapted parenting intervention could be effective across various crisis types. With this concept in mind, the *‘Hope Groups’* parenting program was developed for the Ukrainian war context, as previously reported.^17^ The intervention used key principles for psychosocial support in emergencies to build skills in coping, stress reduction, and self-care, as well as caregiving and violence prevention content based on Parenting for Lifelong Health (PLH) at the University of Oxford, with evidence of effectiveness from 15 randomized trials.^43^ The original *‘Hope Groups’* pre-post study found that improvements in caregiver mental health, positive parenting, and violence prevention were more comprehensive and generally of greater magnitude than those reported in 14 previous studies among displaced populations.^17^ A subsequent *‘Hope Groups’* randomized trial among displaced Ukrainians and those living in active war settings also has examined the effectiveness across all primary and secondary outcomes related to caregiver mental health, reductions in violence against children, parenting practices, and child behavioral issues.^44^ Our adaptation of the *‘Hope Groups’* as an in-home program for next-of-kin caregivers of orphaned children in Colombia in the context of the COVID-19 pandemic, also shows *‘Parenting with Hope’* to be highly effective, with both the pre-post and quasi-experimental findings having high concordance for each outcome. Furthermore, comparisons of findings across diverse populations of bereaved caregivers in Colombia with caregivers experiencing trauma war-linked trauma in Ukraine were fully aligned, thus supporting the external validity of the program. These results suggest the robustness of the *‘Parenting with Hope’/‘Hope Groups’* program and suggest it may be effective in protecting children and families across a wide range of crisis contexts and cultures.

We considered several limitations. First, prompt identification of recently bereaved children and their caregivers posed numerous challenges that required we broaden our recruitment strategies to include rolling recruitment. This limited our ability to assemble a list in advance and randomize potential participants. Our aim of delivering psychosocial support to recently orphaned children and their next-of-kin caregivers in a timely fashion further limited us to using a pre-post and quasi-experimental analytic approach, rather than a randomized trial. Preliminary findings from ongoing randomized trials in Ukraine and Jordan appear to confirm *‘Hope Groups’* as effective. We further noted that one of our primary outcomes, caregiver depression and anxiety, did not meet the assumptions required for use of a Poisson model, and thus required we develop a new methodology for evaluation of likert-style data (see SM). Findings based on this new approach can strengthen rigor, power, and relevance of future treatment effect models. A strength of our approach was a close collaboration with local institutions that ensured we identified as many families as possible in the municipalities where we conducted the intervention. We also selected five municipalities in different regions of the Department to ensure that we capture as many socio-economic and conflict-related differences as possible affecting this region.

In conclusion, both the pre-post and quasi-experimental analyses showed that participation in *‘Parenting for Hope’* was associated with marked reductions in caregiver depression and anxiety and strengthened positive parenting and prevention of violence against children. The promising effectiveness of this intervention is encouraging, given the level of trauma and socio-economic challenges that children and families in Colombia are experiencing. Strong collaboration with local government and academic institutions further ensures sustainability but also contributes to scalability at a national level. Our research suggests that adaptations of this evidence-based psychosocial parenting support and violence prevention program (*‘Parenting with Hope’/‘Hope Groups’)* may be effective and replicable in diverse crisis contexts.

## Data Availability

All data produced in the present study are available upon reasonable request to the authors.

## Contributors statement

Drs Andrés Villaveces, Susan D. Hillis, and Oliver Ratmann guided the conceptualization and investigation, wrote the first draft of the article, guided the writing, and led the reviews and critical revisions of the manuscript; Dr Oliver Ratmann and Ms Sydney Tucker designed and performed statistical analyses, led writing of the Methods section, and prepared all visualizations. Dr. Stiven Arroyo managed data collection in the field, linked data from the Department of Health, and contributed to statistical analyses. Ms Pecsérke contributed to the analysis of data. Sydney Tucker verified the data. Dr Seth Flaxman provided key inputs on the modelling and analytical approach. Ms. Mayerlis Colón and Hilda E. Prías, led the implementation of the intervention in the field and contributed to training personnel. Dr. Blanco led the implementation locally in Sucre and provided critically important intellectual content and contributed to establishing key institutional contacts. Ms. Nicole Baldonado provided inputs on the adaptation of the *‘Hope Groups’* into the *‘Parenting with Hope’* program. All authors approved the final manuscript as submitted and agree to be accountable for all aspects of the work.

## Acknowledgements

The authors would like to thank all the social workers and counselors who participated in the training and provided care to the beneficiaries of this pilot program in the five different municipalities of the Department of Sucre. We are also thankful to the local authorities of the Sincelejo Department of Health for providing access to critical datasets and to the leadership of the Universidad de Sucre for providing space and physical resources to conduct training for this intervention. We thank the administrative personnel at the Imperial College London for expediting several processes during the development of this intervention, the Imperial College Research Computing Service (DOI: 10.14469/hpc/2232) for proving computing resources, and Zulip for free-tier access to the Zulip project management application. Finally, the authors wish to thank Ms. Nicole Baldonado from World Without Orphans for providing key inputs for the Colombian adaptation of the original *‘Hope Groups’* program conducted in Ukraine.

---

**Table S1:** Changes in response items following Parenting with Hope participation among next-of-kin caregivers in Colombia in 2024-2025, in the pre-post analysis.

| Type | Item | Direction of Improvement | Baseline<br>(median, 95% CrI) | Endline<br>(median, 95% CrI) | Difference<br>(Endline vs Baseline)<br>(median, 95% CrI) | Percent change<br>(Endline vs Baseline)<br>(median, 95% CrI) |
| --- | --- | --- | --- | --- | --- | --- |
| **Primary (pre-post analysis)** |  |  |  |  |  |  |
|  | Caregiver Depression and Anxiety: effort<sup>†</sup> | Lower | 49.5%<br>(43.6%, 55.4%) | 6.1%<br>(3.4%, 9.8%) | -43.3%<br>(-49.8%, -36.5%) | -84.7%<br>(-92.5%, -61.4%) |
|  | Caregiver Depression and Anxiety: hopeless<sup>†</sup> | Lower | 57.2%<br>(51.6%, 62.7%) | 2.5%<br>(0.9%, 5.2%) | -54.6%<br>(-60.5%, -48.4%) | -94.5%<br>(-98.3%, -69.5%) |
|  | Caregiver Depression and Anxiety: nervous<sup>†</sup> | Lower | 57.7%<br>(51.8%, 63.6%) | 3.4%<br>(1.4%, 6.4%) | -54.2%<br>(-60.8%, -47.5%) | -94.2%<br>(-97.8%, -88.5%) |
|  | Caregiver Depression and Anxiety: restless<sup>†</sup> | Lower | 56.4%<br>(50.5%, 62.2%) | 4.5%<br>(2.3%, 7.8%) | -51.7%<br>(-58.0%, -45.3%) | -90.0%<br>(-95.7%, -65.2%) |
|  | Caregiver Depression and Anxiety: sad<sup>†</sup> | Lower | 61.0%<br>(55.4%, 66.8%) | 5.4%<br>(2.9%, 9.2%) | -55.4%<br>(-61.6%, -48.8%) | -90.6%<br>(-95.3%, -80.5%) |
|  | Caregiver Depression and Anxiety: worthless<sup>†</sup> | Lower | 7.8%<br>(4.7%, 11.8%) | 1.2%<br>(0.3%, 3.1%) | -6.5%<br>(-10.7%, -3.1%) | -86.7%<br>(-97.0%, -56.4%) |
|  | Caregiver depression<sup>‡</sup> | Lower | 4.0<br>(3.8, 4.2) | 1.0<br>(0.8, 1.2) | -3.0<br>(-3.3, -2.8) | -72.2%<br>(-77.0%, -66.2%) |
|  | Caregiver involvement: child-problems<sup>‡</sup> | Higher | 1.7<br>(1.5, 2.0) | 2.5<br>(2.3, 2.8) | 0.8<br>(0.5, 1.2) | 53.7%<br>(27.3%, 85.6%) |
|  | Caregiver involvement: help-learn<sup>‡</sup> | Higher | 3.5<br>(3.2, 3.8) | 4.2<br>(3.9, 4.5) | 0.7<br>(0.3, 1.1) | 20.2%<br>(7.2%, 35.2%) |
|  | Caregiver positive parenting: play<sup>‡</sup> | Higher | 3.1<br>(2.8, 3.5) | 4.1<br>(3.8, 4.4) | 1.0<br>(0.5, 1.4) | 30.7%<br>(15.3%, 49.0%) |
|  | Caregiver positive parenting: praise<sup>‡</sup> | Higher | 3.3<br>(3.0, 3.5) | 4.5<br>(4.2, 4.7) | 1.2<br>(0.8, 1.6) | 35.9%<br>(24.0%, 52.3%) |
|  | Caregiver exercising nonviolent discipline<sup>‡</sup> | Higher | 2.9<br>(2.6, 3.2) | 3.4<br>(3.1, 3.7) | 0.5<br>(0.0, 0.9) | 15.6%<br>(1.0%, 32.8%) |
|  | Caregiver monitoring: child-safe<sup>‡</sup> | Higher | 5.4<br>(5.2, 5.7) | 6.6<br>(6.4, 6.7) | 1.1<br>(0.8, 1.4) | 20.9%<br>(15.2%, 27.3%) |
|  | Caregiver monitoring: safe-time<sup>‡</sup> | Higher | 5.4<br>(5.1, 5.6) | 5.8<br>(5.6, 6.1) | 0.5<br>(0.1, 0.9) | 8.7%<br>(1.3%, 16.9%) |
|  | Violence against children: ph-punish<sup>‡</sup> | Lower | 1.0<br>(0.8, 1.3) | 0.2<br>(0.2, 0.3) | -0.8<br>(-1.1, -0.6) | -78.4%<br>(-86.1%, -67.5%) |
|  | Violence against children: scream<sup>‡</sup> | Lower | 2.0<br>(1.8, 2.3) | 0.8<br>(0.6, 0.9) | -1.2<br>(-1.5, -1.0) | -61.2%<br>(-68.6%, -51.4%) |
| **Secondary (pre-post analysis)** |  |  |  |  |  |  |
|  | Caregiver self-care<sup>‡</sup> | Higher | 2.2<br>(1.9, 2.5) | 5.2<br>(4.9, 5.4) | 3.0<br>(2.6, 3.4) | 139.5%<br>(107.5%, 178.1%) |
|  | Child behavioural issues: angry<sup>‡</sup> | Lower | 3.1<br>(2.9, 3.4) | 1.0<br>(0.8, 1.2) | -2.1<br>(-2.5, -1.8) | -67.3%<br>(-73.4%, -59.8%) |
|  | Child behavioural issues: no-interest<sup>‡</sup> | Lower | 1.7<br>(1.5, 1.9) | 0.3<br>(0.2, 0.4) | -1.4<br>(-1.7, -1.2) | -81.7%<br>(-87.8%, -72.9%) |
|  | Child behavioural issues: unhappy<sup>‡</sup> | Lower | 3.1<br>(2.8, 3.3) | 0.6<br>(0.5, 0.8) | -2.4<br>(-2.7, -2.1) | -78.4%<br>(-83.1%, -72.0%) |
|  | Caregiver Hope and Resilience<sup>‡</sup> | Higher | 4.6<br>(4.3, 4.9) | 6.5<br>(6.4, 6.7) | 2.0<br>(1.7, 2.3) | 43.8%<br>(34.5%, 54.8%) |

<sup>†</sup> Unit of outcome: frequency of the two categories 'most of the time' and 'all of the time'.
<sup>‡</sup> Unit of outcome: mean days per week.

**Table S2:**
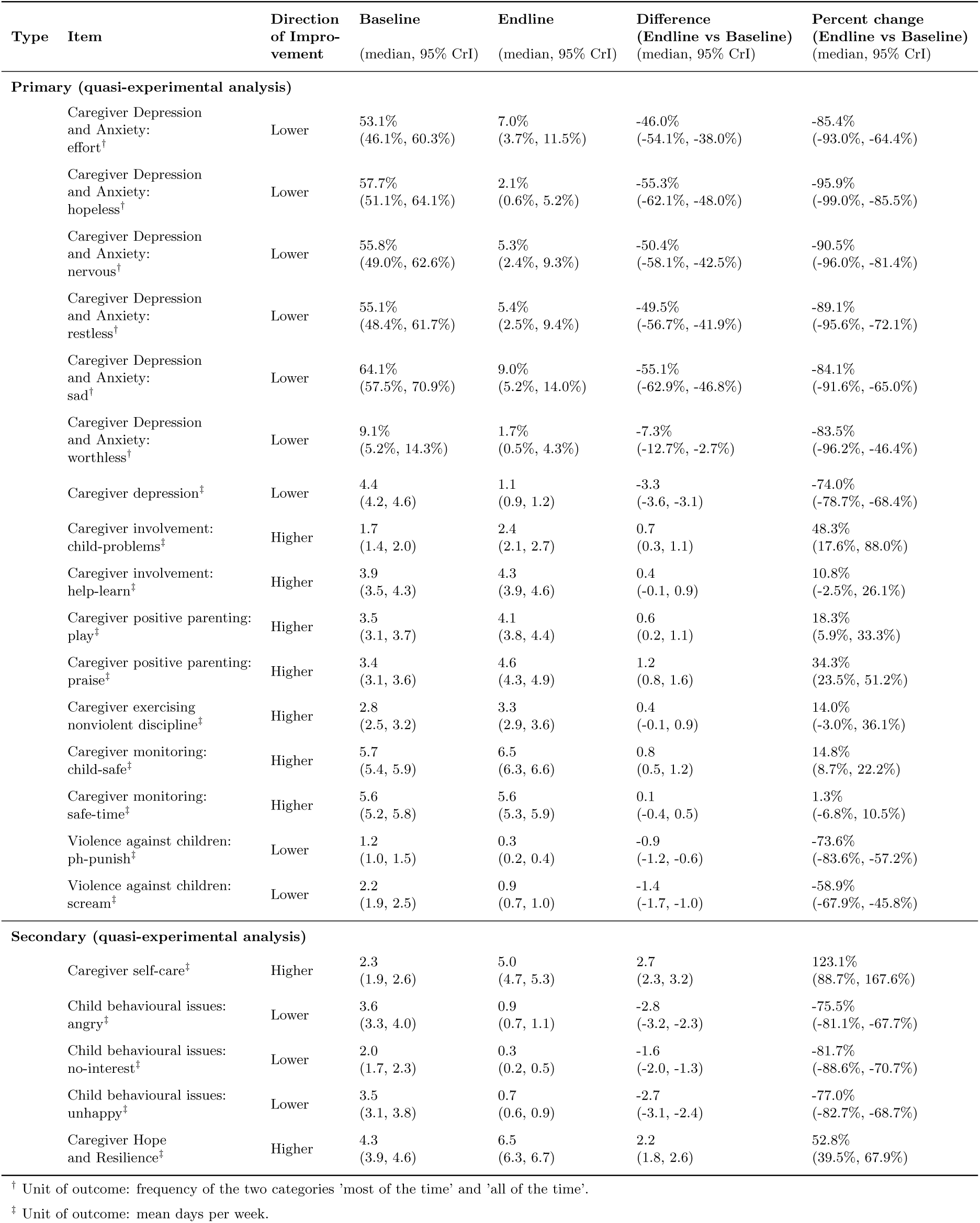
Changes in response items following Parenting with Hope participation among next-of-kin caregivers in Colombia in 2024-2025, in the quasi-experimental analysis.

| Type | Item | Direction of Improvement | Baseline<br>(median, 95% CrI) | Endline<br>(median, 95% CrI) | Difference<br>(Endline vs Baseline)<br>(median, 95% CrI) | Percent change<br>(Endline vs Baseline)<br>(median, 95% CrI) |
| --- | --- | --- | --- | --- | --- | --- |
| **Primary (quasi-experimental analysis)** |  |  |  |  |  |  |
|  | Caregiver Depression and Anxiety: effort<sup>†</sup> | Lower | 53.1%<br>(46.1%, 60.3%) | 7.0%<br>(3.7%, 11.5%) | -46.0%<br>(-54.1%, -38.0%) | -85.4%<br>(-93.0%, -64.4%) |
|  | Caregiver Depression and Anxiety: hopeless<sup>†</sup> | Lower | 57.7%<br>(51.1%, 64.1%) | 2.1%<br>(0.6%, 5.2%) | -55.3%<br>(-62.1%, -48.0%) | -95.9%<br>(-99.0%, -85.5%) |
|  | Caregiver Depression and Anxiety: nervous<sup>†</sup> | Lower | 55.8%<br>(49.0%, 62.6%) | 5.3%<br>(2.4%, 9.3%) | -50.4%<br>(-58.1%, -42.5%) | -90.5%<br>(-96.0%, -81.4%) |
|  | Caregiver Depression and Anxiety: restless<sup>†</sup> | Lower | 55.1%<br>(48.4%, 61.7%) | 5.4%<br>(2.5%, 9.4%) | -49.5%<br>(-56.7%, -41.9%) | -89.1%<br>(-95.6%, -72.1%) |
|  | Caregiver Depression and Anxiety: sad<sup>†</sup> | Lower | 64.1%<br>(57.5%, 70.9%) | 9.0%<br>(5.2%, 14.0%) | -55.1%<br>(-62.9%, -46.8%) | -84.1%<br>(-91.6%, -65.0%) |
|  | Caregiver Depression and Anxiety: worthless<sup>†</sup> | Lower | 9.1%<br>(5.2%, 14.3%) | 1.7%<br>(0.5%, 4.3%) | -7.3%<br>(-12.7%, -2.7%) | -83.5%<br>(-96.2%, -46.4%) |
|  | Caregiver depression<sup>‡</sup> | Lower | 4.4<br>(4.2, 4.6) | 1.1<br>(0.9, 1.2) | -3.3<br>(-3.6, -3.1) | -74.0%<br>(-78.7%, -68.4%) |
|  | Caregiver involvement: child-problems<sup>‡</sup> | Higher | 1.7<br>(1.4, 2.0) | 2.4<br>(2.1, 2.7) | 0.7<br>(0.3, 1.1) | 48.3%<br>(17.6%, 88.0%) |
|  | Caregiver involvement: help-learn<sup>‡</sup> | Higher | 3.9<br>(3.5, 4.3) | 4.3<br>(3.9, 4.6) | 0.4<br>(-0.1, 0.9) | 10.8%<br>(-2.5%, 26.1%) |
|  | Caregiver positive parenting: play<sup>‡</sup> | Higher | 3.5<br>(3.1, 3.7) | 4.1<br>(3.8, 4.4) | 0.6<br>(0.2, 1.1) | 18.3%<br>(5.9%, 33.3%) |
|  | Caregiver positive parenting: praise<sup>‡</sup> | Higher | 3.4<br>(3.1, 3.6) | 4.6<br>(4.3, 4.9) | 1.2<br>(0.8, 1.6) | 34.3%<br>(23.5%, 51.2%) |
|  | Caregiver exercising nonviolent discipline<sup>‡</sup> | Higher | 2.8<br>(2.5, 3.2) | 3.3<br>(2.9, 3.6) | 0.4<br>(-0.1, 0.9) | 14.0%<br>(-3.0%, 36.1%) |
|  | Caregiver monitoring: child-safe<sup>‡</sup> | Higher | 5.7<br>(5.4, 5.9) | 6.5<br>(6.3, 6.6) | 0.8<br>(0.5, 1.2) | 14.8%<br>(8.7%, 22.2%) |
|  | Caregiver monitoring: safe-time<sup>‡</sup> | Higher | 5.6<br>(5.2, 5.8) | 5.6<br>(5.3, 5.9) | 0.1<br>(-0.4, 0.5) | 1.3%<br>(-6.8%, 10.5%) |
|  | Violence against children: ph-punish<sup>‡</sup> | Lower | 1.2<br>(1.0, 1.5) | 0.3<br>(0.2, 0.4) | -0.9<br>(-1.2, -0.6) | -73.6%<br>(-83.6%, -57.2%) |
|  | Violence against children: scream<sup>‡</sup> | Lower | 2.2<br>(1.9, 2.5) | 0.9<br>(0.7, 1.0) | -1.4<br>(-1.7, -1.0) | -58.9%<br>(-67.9%, -45.8%) |
| **Secondary (quasi-experimental analysis)** |  |  |  |  |  |  |
|  | Caregiver self-care<sup>‡</sup> | Higher | 2.3<br>(1.9, 2.6) | 5.0<br>(4.7, 5.3) | 2.7<br>(2.3, 3.2) | 123.1%<br>(88.7%, 167.6%) |
|  | Child behavioural issues: angry<sup>‡</sup> | Lower | 3.6<br>(3.3, 4.0) | 0.9<br>(0.7, 1.1) | -2.8<br>(-3.2, -2.3) | -75.5%<br>(-81.1%, -67.7%) |
|  | Child behavioural issues: no-interest<sup>‡</sup> | Lower | 2.0<br>(1.7, 2.3) | 0.3<br>(0.2, 0.5) | -1.6<br>(-2.0, -1.3) | -81.7%<br>(-88.6%, -70.7%) |
|  | Child behavioural issues: unhappy<sup>‡</sup> | Lower | 3.5<br>(3.1, 3.8) | 0.7<br>(0.6, 0.9) | -2.7<br>(-3.1, -2.4) | -77.0%<br>(-82.7%, -68.7%) |
|  | Caregiver Hope and Resilience<sup>‡</sup> | Higher | 4.3<br>(3.9, 4.6) | 6.5<br>(6.3, 6.7) | 2.2<br>(1.8, 2.6) | 52.8%<br>(39.5%, 67.9%) |

<sup>†</sup> Unit of outcome: frequency of the two categories 'most of the time' and 'all of the time'.
<sup>‡</sup> Unit of outcome: mean days per week.

**Table S3:**
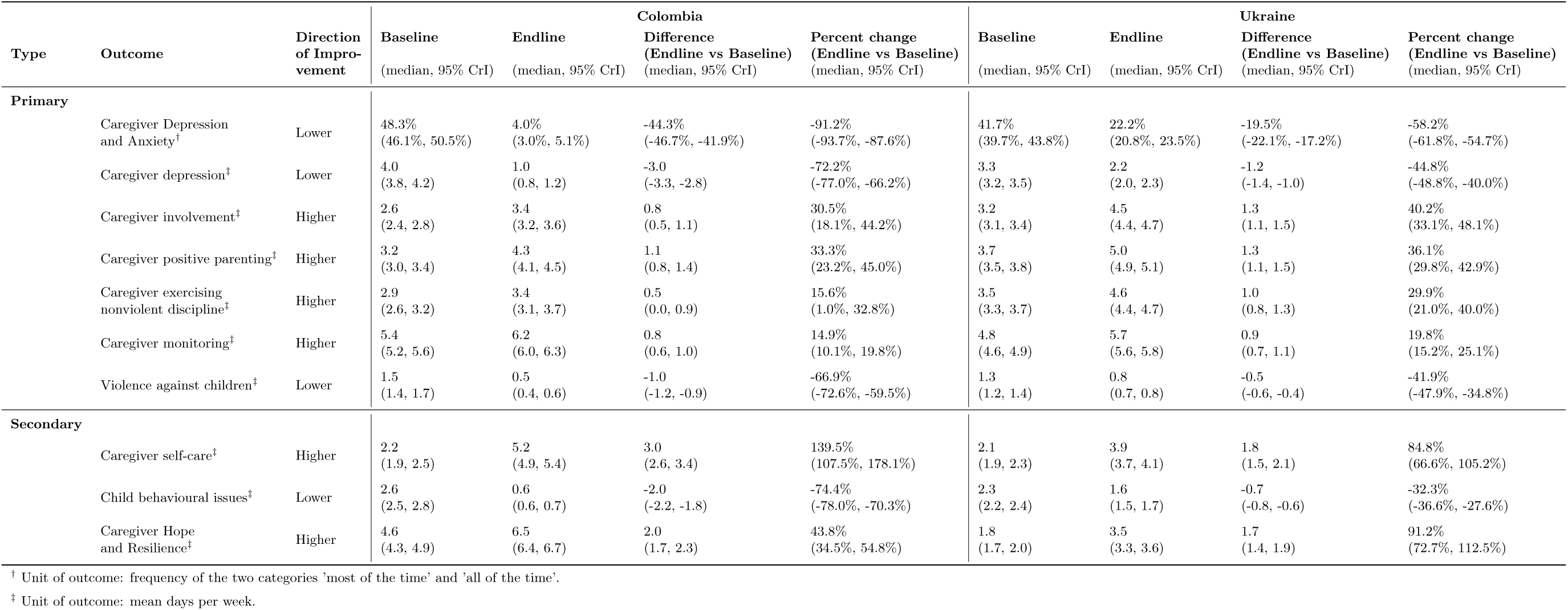
Comparing Parenting with Hope and Hope Groups across global regions and crisis settings: bereaved next-of-kin caregivers in Colombia (2024-2025) versus Ukrainian caregivers amidst war and displacement (2023)

**Supplementary Figure S1:**
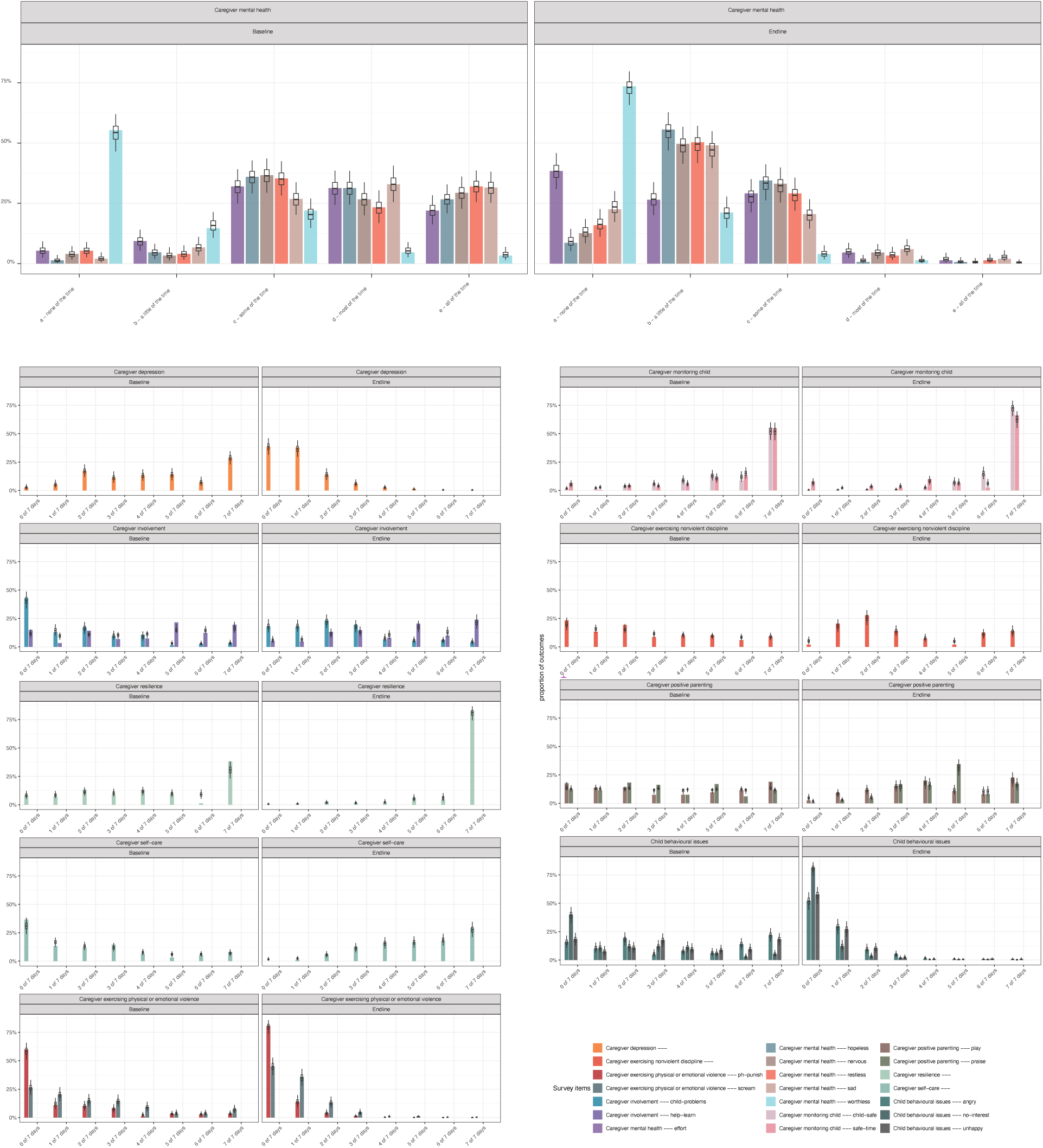
Goodness-of-fit evaluation of the Bayesian partial credit model to all survey items following *‘Parenting with Hope’* participation among next-of-kin caregivers in Colombia (2024-2025).

**Supplementary Figure S2:**
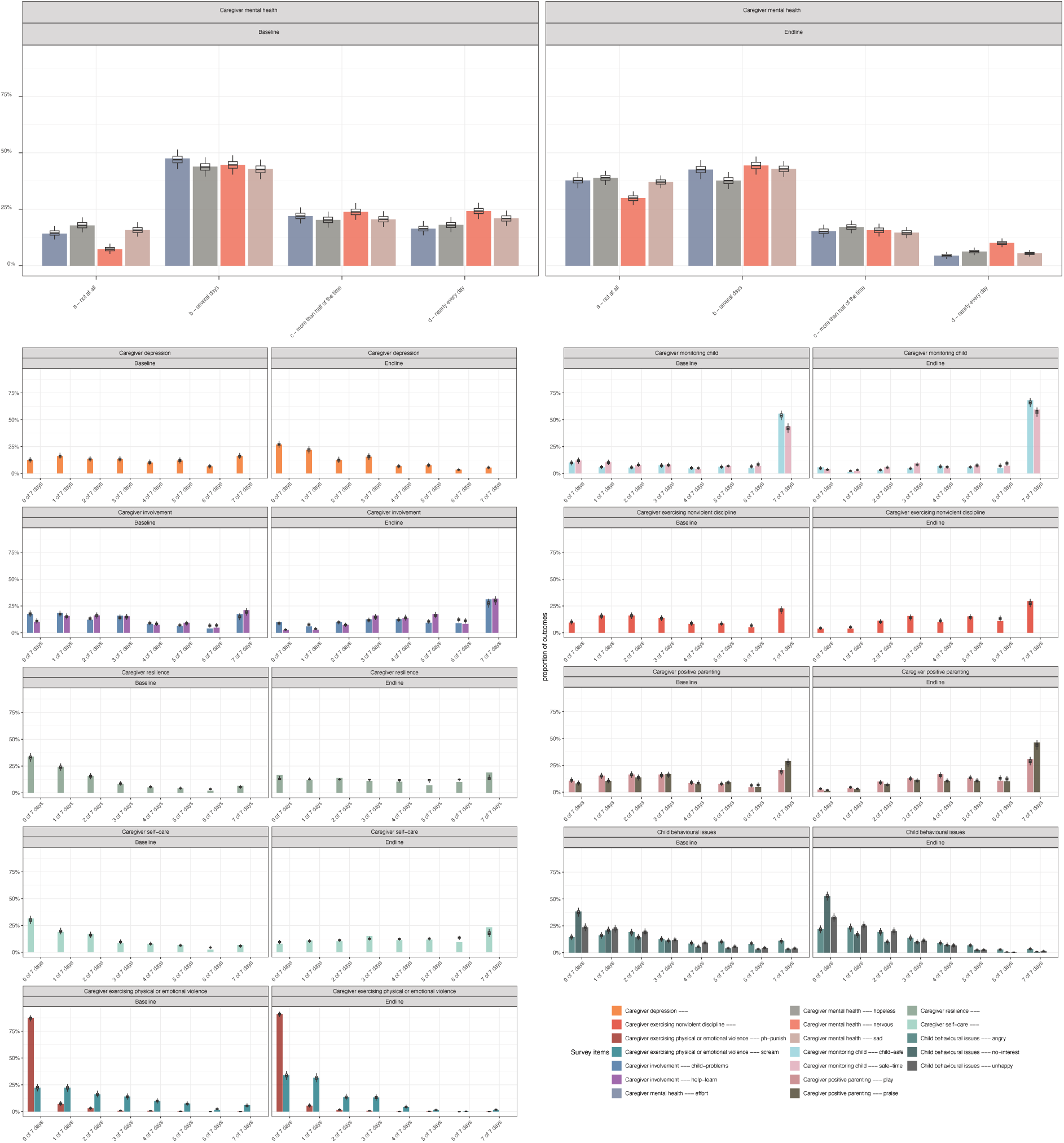
Goodness-of-fit evaluation of the Bayesian partial credit model to all survey items following *‘Hope Group’* participation among next-of-kin caregivers in Ukraine (2023).

**Supplemental Figure S3.**
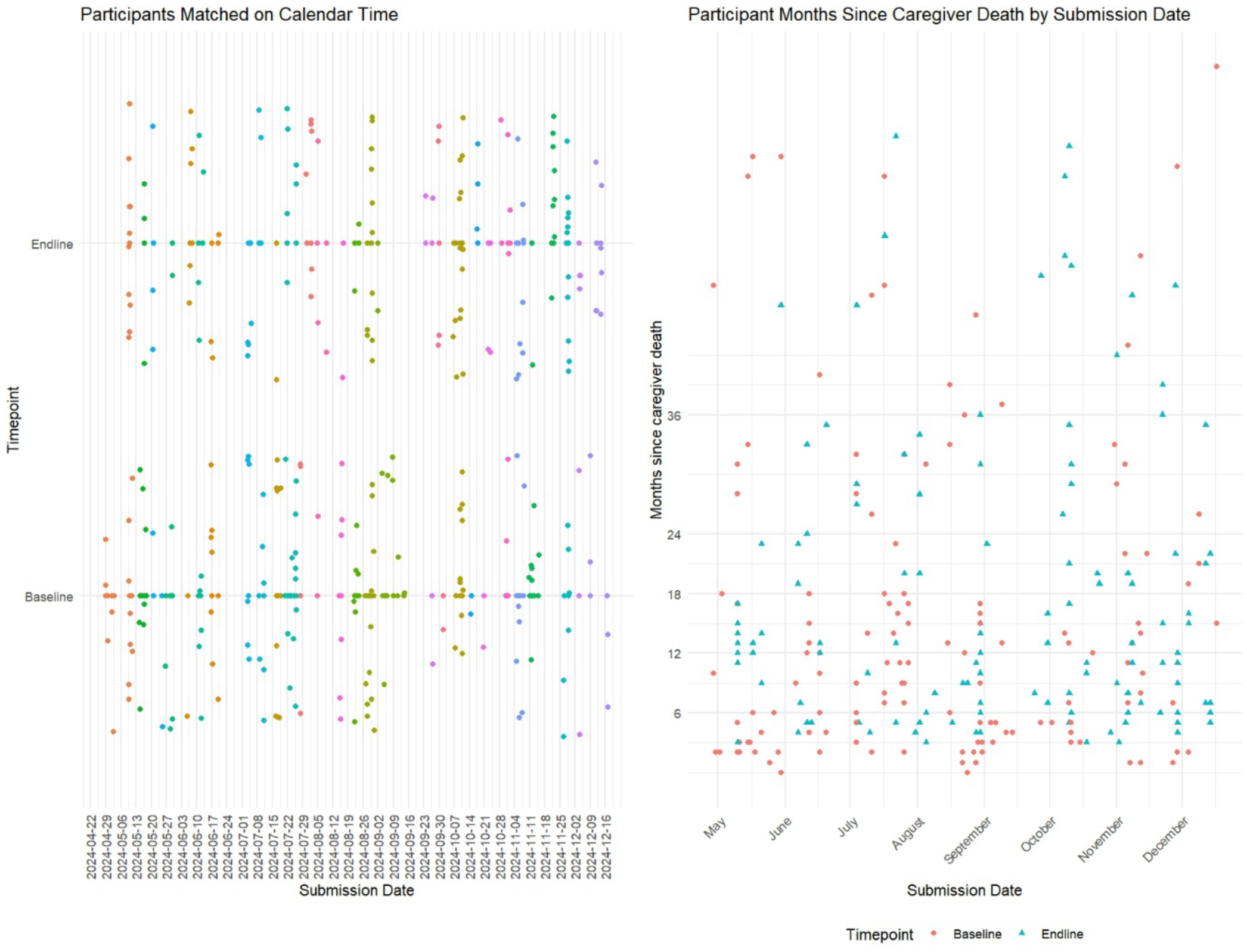
Quasi-Experimental Stepped Entry Matching: Baseline and Endline Data Displayed by Calendar Time and Time Since Caregiver Death.

